# MindWatch: A Smart Cloud-based AI solution for Suicide Ideation Detection leveraging Large Language Models

**DOI:** 10.1101/2023.09.25.23296062

**Authors:** Runa Bhaumik, Vineet Srivastava, Arash Jalali, Shanta Ghosh, Ranganathan Chandrasekharan

## Abstract

Suicide, a serious public health concern affecting millions of individuals worldwide, refers to the intentional act of ending one’s own life. Mental health issues such as depression, frustration, and hopelessness can directly or indirectly influence the emergence of suicidal thoughts. Early identification of these thoughts is crucial for timely diagnosis. In recent years, advances in artificial intelligence (AI) and natural language processing (NLP) have paved the way for revolutionizing mental health support and education. In this proof-of-concept study, we have created MindWatch, a cutting-edge tool that harnesses the power of AI-driven language models to serve as a valuable computer-aided system for the mental health professions to achieve two important goals such as *early symptom detection, and personalized psychoeducation*. We utilized ALBERT and Bio-Clinical BERT language models and fine-tuned them with the Reddit dataset to build the classifiers. We evaluated the performance of bi-LSTM, ALBERT, Bio-Clinical BERT, OpenAI GPT3.5 (via prompt engineering), and an ensembled voting classifier to detect suicide ideation. For personalized psychoeducation, we used the state-of-the-art Llama 2 foundation model leveraging prompt engineering. The tool is developed in the Amazon Web Service environment. All models performed exceptionally well, with accuracy and precision/recall greater than 92%. ALBERT performed better (AUC=.98) compared to the zero-shot classification accuracies obtained from OpenAI GPT3.5 Turbo (ChatGPT) on hidden datasets (AUC=.91). Furthermore, we observed that the inconclusiveness rate of the Llama 2 model is low while tested for few examples. This study emphasizes how transformer models can help provide customized psychoeducation to individuals dealing with mental health issues. By tailoring content to address their unique mental health conditions, treatment choices, and self-help resources, this approach empowers individuals to actively engage in their recovery journey. Additionally, these models have the potential to advance the automated detection of depressive disorders.

## Introduction

The causes of suicide are complicated and can arise from the interaction of multiple factors such as health, environment, and personal history, such as childhood abuse or previous suicide attempts ^1,2^. Additional examples of suicide risk factors include mental disorders, physical illness, substance abuse, domestic violence, bullying, relationship difficulties, and other significant life stressors. Given the complexity of the issue, no single risk factor can reliably predict suicide^3^. Furthermore, the ongoing COVID-19 pandemic has introduced additional challenges to people’s well-being and mental health, stemming from factors such as increased mortality rates, social isolation, and job losses. These circumstances further contribute to the heightened risk of suicide ^4^. The early detection of suicidal thoughts is the key to prevention through health professionals. However, there are several challenges associated with suicide prevention which include social stigma, limited access to professional help, and inadequate training of clinicians. These lead to a new of fragmented professional care 5,6 for patients in accessing and receiving the necessary support they need.

Traditionally, suicide research has relied on structured data (i.e., close-ended) to examine risk factors (e.g., demographics, mental health diagnoses, substance use, social support) and evaluate prevention efforts (e.g., mental health treatment, restricting access to lethal means) ^7^. However, there is growing recognition of the value of unstructured textual information in gaining a deeper understanding of individuals’ experiences and identifying new risk factors. With the growth of digital media, there has been a significant impact on the field of suicide research. Digital media encompasses various online platforms, social networking sites, forums, blogs, and other forms of online communication. These platforms have become important sources of information and insights into individuals’ mental health, including their experiences with suicidal thoughts and behaviours. As these texts are easily accessible, they became valuable resources for research studies that utilize machine learning (ML), deep learning (DL), and natural language processing (NLP) techniques to detect and identify suicidal ideation.

### Challenges with Traditional Machine Learning and Natural Language Processing (NLP) Approaches

Supervised Machine Learning (ML) models such as Logistic Regression, Random Forest, Naïve Bayes, or advanced Natural Language Processing (NLP) models such as LSTM (Long Short-Term Memory) networks, can encounter challenges when capturing sentiments from large social media posts due to several reasons such as noisy and informal language), ambiguity and contextual understanding ^8^, sarcasm and irony^9^, evolving language and neologisms^10^, privacy and ethical concern^11^, labeling and annotation^12^, class imbalance^13^ (De Choudhury et al., 2017).

### State-of-the-Art Artificial Intelligence Models

The State-of-the-art AI models based on the transformers have transformed the NLP landscape in several ways:

#### 1 Representational learning

Uses deep learning techniques to automatically learn hierarchical and contextual representations of language.

#### 2 Scale and Size

They are massive in scale with billions of parameters.

#### 3 Contextual Understanding

They consider the surrounding words when predicting the meaning of a word, making them highly context-aware.

#### 4 Transfer learning

Models are trained on massive amounts of text data and then fine-tuned for specific tasks.

#### 5 Multimodal Capabilities

Advanced AI models can process and generate text, images, and audio, and even combine modalities, enabling applications in image captioning, speech recognition, and more.

#### 6 Few-shot and Zero-shot Learning

Advanced models like GPT-3 can perform tasks with very few examples or even zero examples, showcasing their ability to generalize and adapt to new tasks without extensive training data.

#### 7 Ethical Considerations and Bias

Large AI models have raised concerns about bias and ethical issues due to the data they are trained on and their capacity to generate human-like text. Addressing these challenges is a priority in the field.

These advantages make the advanced AI models highly versatile and effective in various natural language understanding and generation tasks by overcoming the issue of training models with large, labeled data. Many researchers leveraged transformer-based pre-trained language representation models in mental health research, including BERT ^14^, DistilBERT^15^, Roberta^16^, ALBERT^17^, BioClinical BERT for clinical notes ^18^, XLNET ^13^, and GPT model ^19^.

In this research, we evaluated the performance of ALBERT, Bio-Clinical BERT, Bi-LSTM and a voting classifier for suicide ideation detection. We also compared the classifier performances with GPT turbo 3.5 model (ChatGPT).

In our pursuit of personalized psychoeducation and uncovering the causes of depression, we harnessed the capabilities of Llama 2 foundation models within the Amazon SageMaker Studio environment. Llama 2, a robust and efficient large language model (LLM), exhibits the capacity to generate text and code in response to prompts, akin to other chatbot-like systems. Our evaluation of the Llama 2 model was based on a limited subset of samples from the evaluation dataset.

## Data collection and Methods

We used Reddit dataset that contains 2,32,000 posts marked as suicidal or non-suicidal. We used 200,000 posts for building the models. The dataset is divided into 80% training and 20% testing. The remaining posts were kept (32000 posts) for evaluation.

We utilized ALBERT and Bio-Clinical BERT language models and fine-tune them with the above Reddit dataset to build the classifiers. We evaluated the performance of bi-LSTM, ALBERT, Bio-Clinical BERT, and an ensembled voting classifier to detect suicide ideation. The hyperparameters for ALBERT and Bio-Clinical BERT were selected based on the default values commonly used in similar studies. The final hyperparameters used in our experiments were Learning Rate=.01, Batch Size = 128, and Maximum Sequence Length = 512.

To offer tailored diagnosis recommendations based on patient notes and text, we employed the concept of search and retrieval-augmented generation (RAG) as depicted in Figure 2. This approach leveraged the FAISS (Facebook AI Similarity Search) algorithm for retrieving similar documents, utilized sentence transformers for generating embeddings, and incorporated AWS foundation models like LLaMa2-7b-chat for augmentation. This method has been evaluated solely on few samples from evaluation set. For evaluation, we report four widely used metrics in this task, accuracy, precision, recall, and AUC score to provide a comprehensive and informative evaluation of the performance of the classification models.

**Figure 1:**
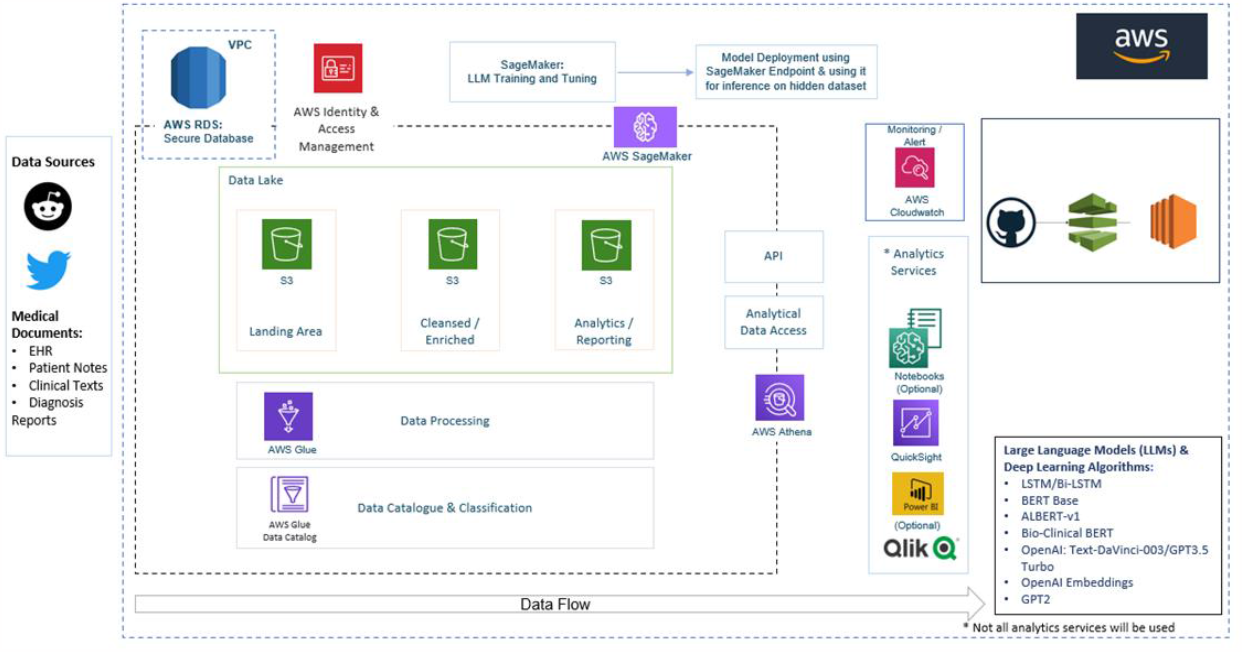
AWS Architecture Diagram

**Figure 2:**
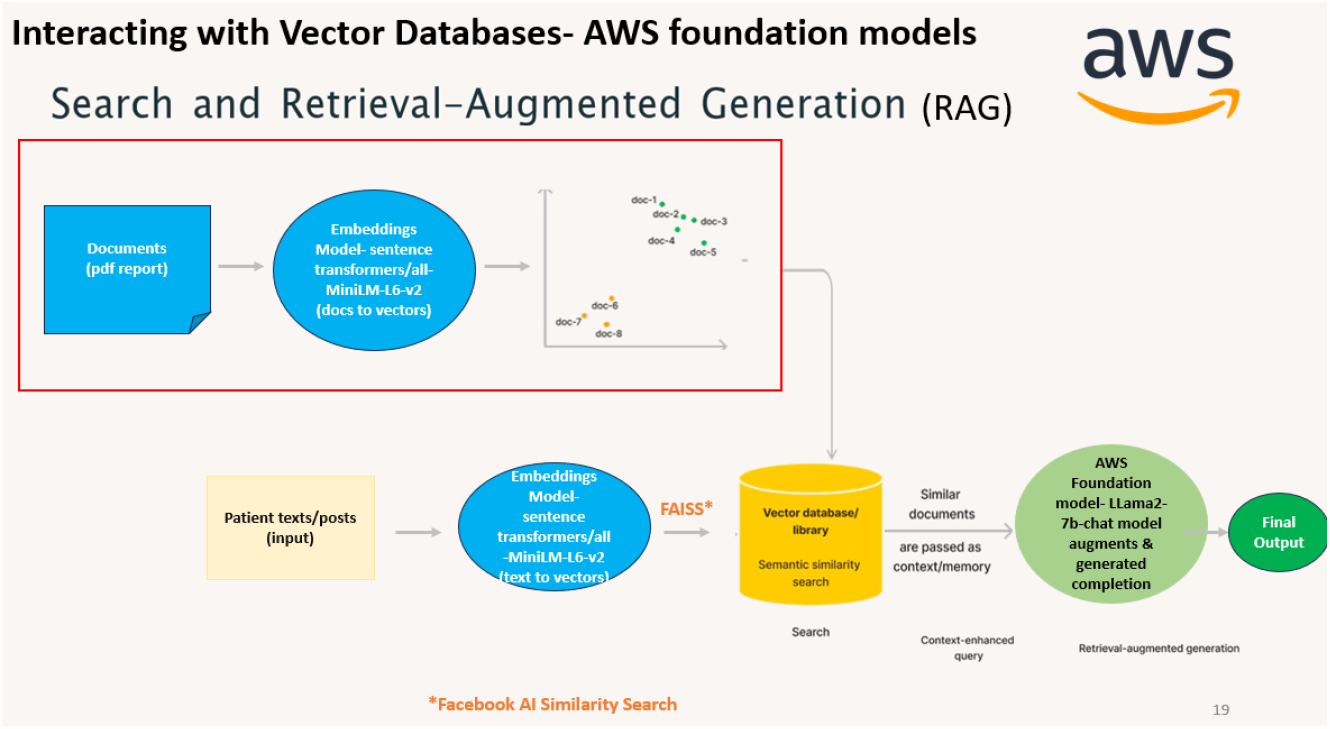
RAG for tailored recommendations/diagnosis using AWS founndation models

**Figure 3:**
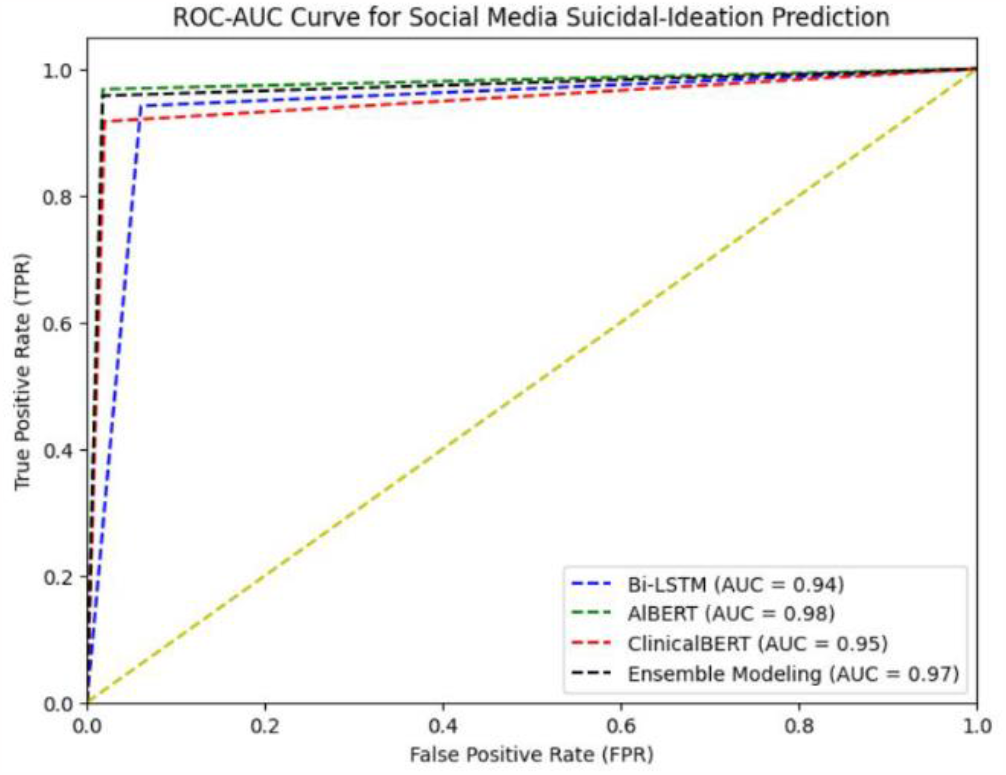
Social Media Suicidal-Ideation Prediction Results using Custom LLMs

### AWS Solution Overview

The [Amazon SageMaker Studio](https://docs.aws.amazon.com/sagemaker/latest/dg/studio.html) is the integrated development environment (IDE) within [Amazon SageMaker](https://aws.amazon.com/sagemaker/) that provides us with all the ML features that we need in a single pane of glass. Training and fine-tuning Deep Learning (DL) and Large language models (LLMs) like bi-LSTM, BERT, GPT, and other advanced architectures often require substantial computational resources, given the large number of parameters they have. We utilized the SageMaker training instances, like ml.m5.16x large, and ml.g5.48xlarge along with the **Reddit dataset** that contains **2**,**32**,**000** labeled records marked as suicidal or non-suicidal.

Below is the architecture of the AWS ecosystem we used for training and plan to further use for the deployment of the tool.

The workflow includes the following steps:

1. We designed a Data Lake Architecture using [AWS Simple Storage Service (S3)](https://aws.amazon.com/s3/). The raw training data was initially ingested in the staging (Bronze) S3 bucket.
2. [AWS Glue](https://aws.amazon.com/glue/) services including [Glue Crawlers](https://docs.aws.amazon.com/glue/latest/dg/add-crawler.html), [Glue ETL Jobs](https://docs.aws.amazon.com/glue/latest/dg/aws-glue-api-jobs-job.html) and [Glue Data Catalog](https://docs.aws.amazon.com/prescriptive-guidance/latest/serverless-etl-aws-glue/aws-glue-data-catalog.html) was used for converting the raw csv data to parquet format and the same was stored in silver S3 bucket, followed by pre-processing & cleaning of raw texts/posts. The pre-processed and cleaned texts/dataset was stored in S3 Gold bucket-enriched data, ready for consumption for model training/visualization.
3. The [AWS Athena](https://aws.amazon.com/athena/) was used to perform SQL Queries on cleaned Glue Database tables and the same was used by [AWS QuickSight](https://aws.amazon.com/quicksight/) Service for visualizations and exploring word-counts.
4. Finally, the state-of-art models were trained/fine-tuned on Amazon SageMaker studio by consuming the final (Gold) S3 bucket data.
5. The model artifacts, after training/fine-tuning, such as model weights, tokenizers, config files, etc. was saved in another S3 bucket to make use for inference on hidden dataset.
6. The fine-tuned BERT models-ALBERT and Bio-Clinical BERT artifacts are also uploaded on [hugging face portal](https://huggingface.co/vineet1409) which makes it easier to use while using it for developing an AI application or tool.
7. We also wanted to provide customized diagnosis recommendations based on the patient’s notes/texts and hence we used the search and retrieval-augmented generation (RAG^20^) concept (Figure 2) using FAISS (Facebook AI Similarity Search) algorithm for similar document retrieval, sentence transformers to create embeddings and AWS foundation models-LLaMa2-7b-chat for augmentation.

## Results

### Classification Models

To train a classification model on Reddit dataset, we employed two pretrained transformer models ALBERT and Bio-Clinical BERT, and compared the results with Bi-LSTM, and ensembled classifier. The performances of these models on the aforementioned dataset are presented in Table 1.

**Table 1:**

| AI Language models | Accuracy | Precision | Recall |
| --- | --- | --- | --- |
| Bi-LSTM | 0.941 | 0.938 | 0.942 |
| ALBERT | 0.975 | 0.981 | 0.968 |
| Bio-Clinical BERT | 0.948 | 0.978 | 0.917 |
| Ensembled (Voting Classifier) | 0.970 | 0.982 | 0.958 |

All the four models perform exceptionally well, with accuracy and precision/recall greater than 92%. However, ALBERT has been performing better than all the other custom trained/fine-tuned models. Even ALBERT performs better (AUC=.98) compared to the zero-shot classification accuracies obtained from OpenAI GPT3.5 Turbo (ChatGPT) on hidden datasets (AUC=.91) as depicted in Figure 4.

**Figure 4:**
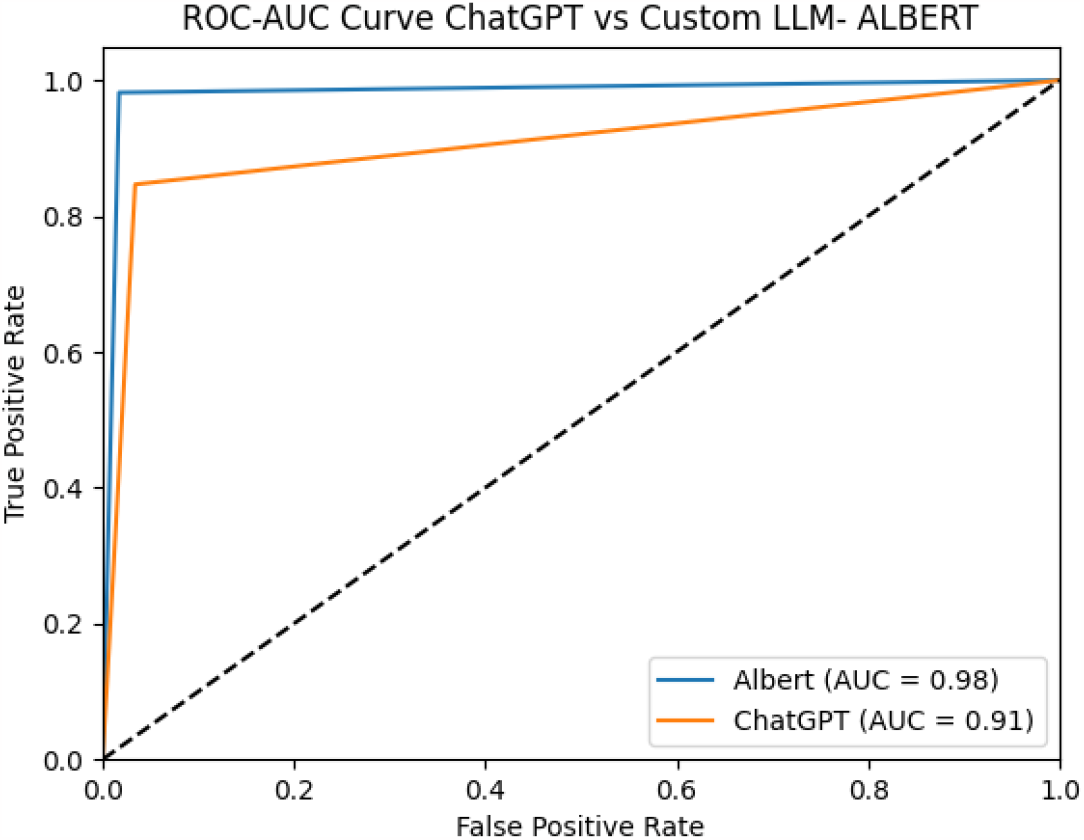
OpenAI ChatGPT vs Custom LLM-ALBERT on hiddent 5000 records

### Generating Healthcare diagnosis and treatment planning

We implemented RAG, an AI framework designed to enhance the quality of responses generated by large language models (LLMs) by enriching them with external knowledge sources. RAG consists of two key phases: retrieval and content generation.

In the retrieval phase, algorithms search for and extract relevant information snippets from a curated set of external documents. These snippets are then added to the user’s prompt, creating an augmented input that is subsequently presented to the language model.

In the generative phase, the LLM utilizes both the augmented input and its internal understanding of training data to craft a tailored and informative response to the user’s query. This response can include links to the sources of the information for further reference by patients or doctors. Figure 5 depicts a snapshot of RAG results.

**Figure 5:**
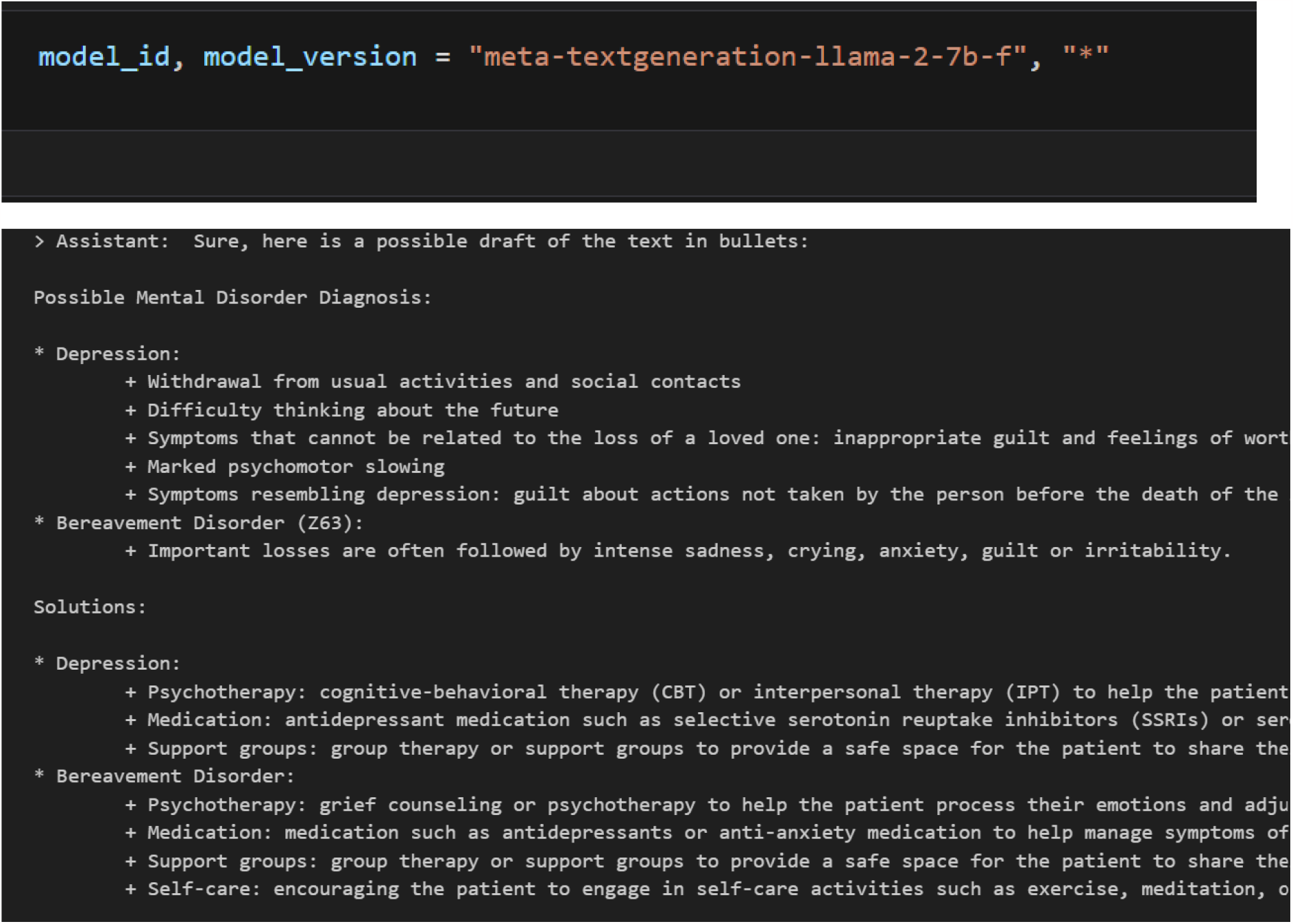
LlaMA2 as assistant for text augmentation

### Key Features of AI Tool developed

#### Robust Integration

Seamlessly combines Llama2 with Custom Language Models (LLMs) to offer a comprehensive and dependable solution. It provides prescriptions or potential diagnostic insights based on symptom analysis from text or posts.

#### Physician Customization

Physicians have the flexibility to personalize prescriptions or diagnostic reports recommended by Custom LLMs (including ALBERT and Bio-Clinical BERT). They can achieve this by uploading their specific documents directly through the web tool.

#### Efficient Batch Analysis

Capable of analyzing and making predictions from single files or large batches of files, streamlining the process for efficiency.

#### Summarized Diagnosis Reports

Generates concise and informative diagnosis reports, simplifying the understanding of complex medical information. Additionally, it provides valuable suggestions.

#### Identification of Depression Causes

Goes beyond surface symptoms to detect the underlying factors contributing to depression, enhancing the overall diagnostic process.

A demo version of MindWatch tool has been recorded and uploaded to Google drive^21^.

#### Future works and Potential

To ensure the effectiveness and fairness of suicidedetection using ALBERT and ChatGPT, it is vital to address biases and generalization issues. Conversational models such as ChatGPT are trained on vast amounts of text data, which may contain biases. Future research should focus on developing bias mitigation techniques to prevent the model from perpetuating harmful stereotypes or stigmatizing individuals. Additionally, efforts should be made to enhance the generalization capabilities of the model by training it on diverse datasets encompassing various demographics, cultures, and languages.This will enable the model to better understand and identify suicidal ideation across different populations.

## Conclusion

In conclusion, the AI application, powered by cutting-edge AI language models and an AWS infrastructure, offers a groundbreaking solution for detecting suicidal posts on social media. By accurately identifying individuals at risk of suicide, we can intervene promptly and provide timely support, potentially saving lives. The integration of Custom LLMs combined with Llama2 and hugging face embeddings, ensures high-performance and comprehensive detection capabilities. Through continuous refinement and evaluation, the solution can contribute to a safer and more supportive online environment, fostering mental well-being in our communities.

It is to be noted that text classification related to mental disorders should not be considered a replacement for the professional diagnosis provided by healthcare practitioners. Instead, it serves as a valuable computer-aided system with several key functions such as early symptom detection, personalized psychoeducation, and understanding the causes. It is also crucial to carefully evaluate the use of large language models in such settings to better appreciate their potential and limitations.

## Institutional Review Board Statement

Not applicable.

## Informed Consent Statement

Not applicable.

## Data Availability Statement

Authors can confirm that all relevant data are available upon request.

